# Patient preferences for features associated with leadless versus conventional transvenous cardiac pacemakers

**DOI:** 10.1101/2024.04.19.24306110

**Authors:** Shelby D. Reed, Jui-Chen Yang, Matthew J. Wallace, Jessie Sutphin, F. Reed Johnson, Semra Ozdemir, Stephanie Delgado, Scott Goates, Nicole Harbert, Monica Lo, Bharath Rajagopalan, James E. Ip, Sana M. Al-Khatib

## Abstract

**Background:** Regulatory approval of the first dual-chamber leadless pacemaker (PM) system provides patients an alternative to conventional transvenous pacemakers.

**Objective:** To quantify patients’ preferences for pacemaker features.

**Methods:** Patients with a de-novo PM indication were recruited from 7 US sites to complete a discrete-choice experiment (DCE) survey. Patients chose between pairs of experimentally designed, hypothetical PMs that varied according to PM type (removable leadless, non-removable leadless, conventional transvenous); battery life (5, 8, 12, 15 years); time since regulatory approval (2, 10 years); discomfort for 6 months (none, discomfort); complication risk and infection risk (1%, 5%, 10%/20% for each). Patients with a de-novo pacemaker indication were recruited to complete a web-based survey from seven US sites between May 11, 2022 to May 24, 2023.

**Results:** Choice data from 117 patients indicated that complication risks and infection risks were the most influential. On average, patients preferred removable leadless pacemakers over both non-removable leadless pacemakers (*p*=0.001) and conventional transvenous pacemakers (*p*=0.031). However, latent-class analysis revealed two distinct preference classes. One class preferred leadless pacemakers (50.5%) and the other class preferred conventional transvenous pacemakers (49.5%). The conventional PM class prioritized pacemakers with ten rather than two years since regulatory approval (*p*<0.001) whereas the leadless PM class was insensitive to years since regulatory approval (*p*=0.83). All else equal, patients would accept maximum risks of complications or infections ranging about 5% to 18% to receive their preferred pacemaker type.

**Conclusion:** Latent-class analysis revealed strong patient preferences for the type of PM, with a nearly equal split between recent leadless PM technology and conventional transvenous PMs.

These findings can inform shared decision making between healthcare providers and patients.

## Background

Pacemakers have been used to treat bradyarrhythmias since the 1950s. Approximately 200,000 pacemakers are implanted each year in the US, and this number has been steadily increasing due to an aging population and expanded indications for pacemakers.^1^ Years of real-word evidence support the improved physical functioning and quality of life associated with transvenous pacemakers. ^2^, ^3^ However, these devices are associated with serious adverse events, such as pocket- and lead-related complications and infections.^4,5^ Patients with conventional transvenous pacemakers also report chest discomfort, physical restriction, and cosmetic concerns.^6^ Over time, transvenous pacemakers have undergone significant advancements, including longer battery life, higher quality pacemaker leads, and smaller generator size.

More recently, leadless pacemakers have emerged as an alternative to conventional transvenous pacemakers, offering certain advantages while also presenting unique risks. ^7^ In contrast to transvenous pacemakers, leadless pacemakers are self-contained pulse generators with built-in batteries that are directly implanted in the heart chambers. Their notable advantage is the absence of leads or a pulse generator pocket, thereby eliminating complications related to components of conventional transvenous pacemakers.^8,9^ Because leadless pacemakers are directly implanted in the heart, they are imperceptible and reduce patient discomfort and scarring. In addition, leadless pacemakers minimize the need for post-surgery activity restrictions, which are required to prevent lead dislodgement after conventional transvenous pacemaker implantation. Two recent meta-analyses reported that, compared with conventional transvenous pacemakers, leadless pacemakers have lower risks of complications including re-intervention, device dislodgment, and pneumothorax.^10,11^ However, both meta-analyses revealed a higher risk of pericardial effusion with leadless pacemakers versus conventional transvenous pacemakers.^10,11^ Other drawbacks of leadless pacemakers include a smaller battery and potentially shorter battery life, necessitating more frequent replacements than with conventional transvenous pacemakers. Given different advantages and disadvantages of leadless and conventional transvenous pacemakers, patients could have varying preferences for pacemaker types.

### Objective

The objective of this study was to quantify patient preferences for device-associated risks and design features that differ between leadless and conventional transvenous pacemakers and the tradeoffs patients would accept to receive a pacemaker with desirable features.

## Methods

A discrete-choice experiment (DCE) was designed to elicit patient preferences regarding different pacemaker features (NCT05327101). DCE was selected as a method that is noted for its conceptual rigor and statistical advantages to other preference-elicitation methods.^12,13^ DCEs present series of constructed treatment options, each defined by a set of features or attributes. Each DCE question in this study asked patients to evaluate a pair of pacemakers wherein the attribute levels shown for each varied according to an experimental design with known statistical properties. The relative preference weights for pacemaker features were quantified based on patients’ pacemaker choices. These weights were then used to calculate maximum-acceptable risks (MARs), which indicate how much risk patients would tolerate to receive a pacemaker with their preferred features.

To identify relevant attributes, we reviewed published literature and product information from manufacturers of conventional transvenous and leadless pacemakers. The final selection of attributes was made collaboratively by a team of clinical researchers, electrophysiologists, methods experts, and four patient representatives by prioritizing features that were deemed important by patients and that characterize different types of pacemakers. The final attributes (and levels) included: pacemaker type (pacemaker with leads, removable leadless pacemaker, or non-removable leadless pacemaker); years of battery life (5, 8, 12, or 15 years); years since government approval (2 or 10 years); discomfort where the device was inserted (groin for leadless pacemaker, chest for transvenous pacemaker) for a duration of six months (discomfort or no discomfort)^14^; risk of a complication requiring an operation and 7-night hospital stay (1, 5, or 10/20%); and risk of infection requiring device removal and four weeks of antibiotics (1, 5, or 10/20%). For the last two attributes related to risk, participants were randomly assigned either a 10% or 20% as the highest risk level as part of a scope test (supplemental materials).

Pretest interviews were conducted with ten patients who either currently had a pacemaker or had an indication for a pacemaker to assess the appropriateness and understandability of the survey content using a think-aloud protocol in which participants were asked to read the survey instrument aloud and share their thoughts regarding the information and questions presented. In addition, interviewers evaluated participants’ understanding of survey information and graphics and varied risk levels to assess participants’ willingness to accept tradeoffs among pacemaker attributes and to check for internal consistency.

The survey instrument incorporated questions pertaining to patient-reported information on personal health and sociodemographic characteristics, including self-reported gender, descriptions of pacemaker attributes, a graphical tutorial illustrating risks, practice questions to familiarize participants with the DCE question format, and 11 comprehension questions designed to test and reinforce educational material required to complete the survey (supplemental materials). Patient preferences for pacemaker features were elicited using eight DCE questions, with an option for participants to complete four additional DCE questions. The survey was programmed for web-based administration using Lighthouse Studio (Sawtooth Software Provo, UT). The pacemaker profiles shown in the DCE questions were governed by two D-optimal experimental designs generated in SAS (SAS Version 9.4, Cary, NC). ^15, 16^ The first design included 48 questions that were divided into six blocks of eight questions each. To generate the additional four questions, another 24-question design was generated and divided into six blocks of four questions each. Participants were randomized to complete one block from each design.

### Study Setting and Sample

The study enrolled adult patients who were referred to Aveir DR i2i investigational device exemption (IDE) clinical trial sites for pacemaker evaluation for a de-novo pacemaker.^17^ Eligible patients had to be ≥ 18 years old, residents of the United States, able to read English, and able to use a tablet or computer to complete the survey. To minimize selection bias, patients were asked to complete the survey before being approached about their interest in enrolling in the IDE study and receiving information about dual-chamber pacemakers. After recruitment was complete for the IDE study, sites continued to invite patients referred for evaluation for a pacemaker to complete the preference survey. All participants provided informed consent before initiating the online survey (Duke Health IRB Protocol 00109587).

### Statistical Analysis

Descriptive statistics were used to summarize participants’ sociodemographic characteristics. Responses to the DCE questions were analyzed using a random-parameters logit (RPL) model where pacemaker features, serving as independent variables, were effect-coded. Effect-coded parameters represent relative coefficients, or preference weights, that sum to zero across levels for each attribute. Differences in preference weights among levels within each pacemaker feature indicate relative strength of preference.

Significant random effects in RPL models indicate the presence of preference heterogeneity. To investigate further, latent-class analysis (LCA) was applied, fitting data to models with up to four latent classes. Model-fit statistics and qualitative differences in preference patterns across latent classes were considered to determine the final number of classes. Fractional multinomial logit regression was used to test for associations between the participant characteristics and latent-class membership. Prespecified covariates included gender, age, body mass index, history of major surgery in the past five years, and level of physical activity.

For each latent class, the maximum risk of a complication (or infection) participants would accept for desirable improvements in pacemaker features was calculated using preference weights from the LCA model. ^18^ Linear approximations for the risk slopes were applied in maximum-acceptable risks calculations, inclusive of the 1% baseline (i.e. lowest) risk levels shown. The Krinsky-Robb method was used to calculate 95% confidence intervals.^19^

## Results

One-hundred twenty-nine patients across seven study sites initiated the survey as of June 13, 2023. Seven did not proceed past the online consent form, and five discontinued the survey before completing it, leaving 117 patients who completed the survey. The mean age was 67.3 years, 94% were White, and about 42% were female (**Table 1**). Fifteen participants (12.8%) were subsequently enrolled in the Aveir DR i2i IDE study. The remaining participants either declined participation in the IDE study (n= 9, 7.7%) or were enrolled after the IDE study completed recruitment (n= 93, 79.5%). The median time spent completing the survey was 32.6 minutes. Participants correctly answered a mean of 9.0 (SD, 2.2) of 11 comprehension questions. About two-thirds (65%) of the participants answered 12 DCE questions; the remainder answered eight.

**Table 1.**
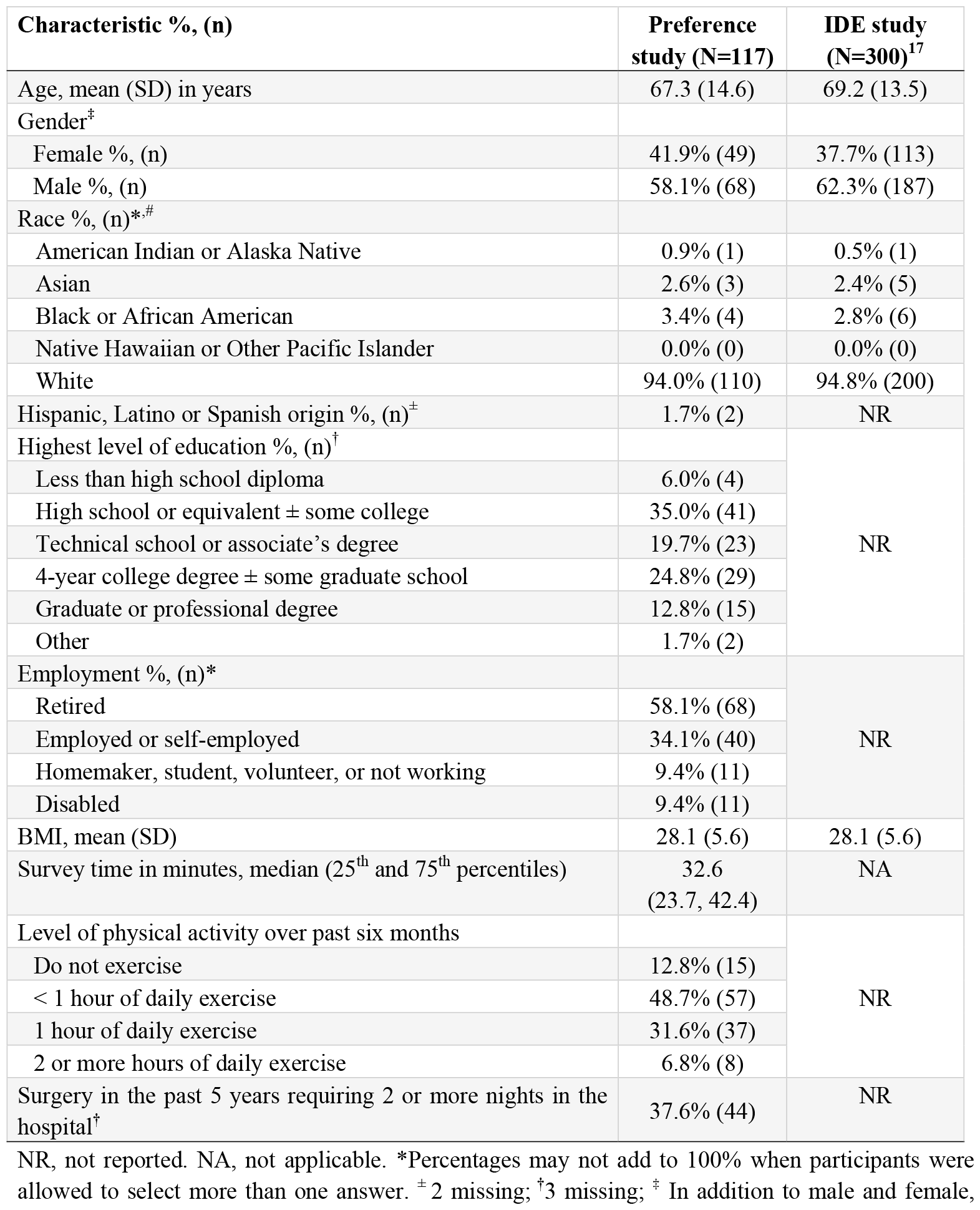
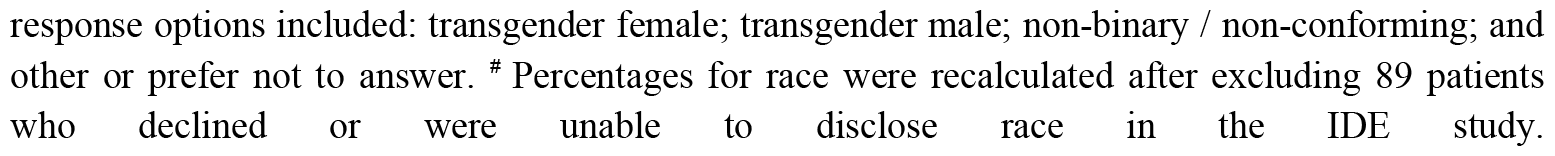
Participant characteristics.

| Characteristic %, (n) | Preference study (N=117) | IDE study (N=300)<sup>17</sup> |
| --- | --- | --- |
| Age, mean (SD) in years | 67.3 (14.6) | 69.2 (13.5) |
| Gender<sup>‡</sup> |  |  |
| Female %, (n) | 41.9% (49) | 37.7% (113) |
| Male %, (n) | 58.1% (68) | 62.3% (187) |
| Race %, (n)<sup>*,#</sup> |  |  |
| American Indian or Alaska Native | 0.9% (1) | 0.5% (1) |
| Asian | 2.6% (3) | 2.4% (5) |
| Black or African American | 3.4% (4) | 2.8% (6) |
| Native Hawaiian or Other Pacific Islander | 0.0% (0) | 0.0% (0) |
| White | 94.0% (110) | 94.8% (200) |
| Hispanic, Latino or Spanish origin %, (n)<sup>±</sup> | 1.7% (2) | NR |
| Highest level of education %, (n)<sup>†</sup> |  | NR |
| Less than high school diploma | 6.0% (4) |  |
| High school or equivalent ± some college | 35.0% (41) |  |
| Technical school or associate's degree | 19.7% (23) |  |
| 4-year college degree ± some graduate school | 24.8% (29) |  |
| Graduate or professional degree | 12.8% (15) |  |
| Other | 1.7% (2) |  |
| Employment %, (n)<sup>*</sup> |  | NR |
| Retired | 58.1% (68) |  |
| Employed or self-employed | 34.1% (40) |  |
| Homemaker, student, volunteer, or not working | 9.4% (11) |  |
| Disabled | 9.4% (11) |  |
| BMI, mean (SD) | 28.1 (5.6) | 28.1 (5.6) |
| Survey time in minutes, median (25<sup>th</sup> and 75<sup>th</sup> percentiles) | 32.6<br>(23.7, 42.4) | NA |
| Level of physical activity over past six months |  | NR |
| Do not exercise | 12.8% (15) |  |
| < 1 hour of daily exercise | 48.7% (57) |  |
| 1 hour of daily exercise | 31.6% (37) |  |
| 2 or more hours of daily exercise | 6.8% (8) |  |
| Surgery in the past 5 years requiring 2 or more nights in the hospital<sup>†</sup> | 37.6% (44) | NR |

NR, not reported. NA, not applicable. \*Percentages may not add to 100% when participants were allowed to select more than one answer. <sup>±</sup> 2 missing; <sup>†</sup>3 missing; <sup>‡</sup> In addition to male and female,
response options included: transgender female; transgender male; non-binary / non-conforming; and other or prefer not to answer. # Percentages for race were recalculated after excluding 89 patients who declined or were unable to disclose race in the IDE study.

Preference weights from the RPL model indicated well-ordered preferences, where superior attribute levels had higher preference weights and inferior levels had lower preference weights (**Figure 2, Table S1**). Across the study sample, there was a statistically significant preference for removable leadless pacemakers over both non-removable leadless pacemakers and conventional transvenous pacemakers (*p*=0.001 and *p*=0.031, respectively). There was no significant difference in preferences between transvenous pacemakers and non-removable leadless pacemakers (*p*=0.23). Random effects for pacemaker type, battery life, time since regulatory approval, and risks (bottom of **Table S1**) were statistically significant (*p*<0.05 for each), indicating preference heterogeneity across participants.

**Figure 1.**
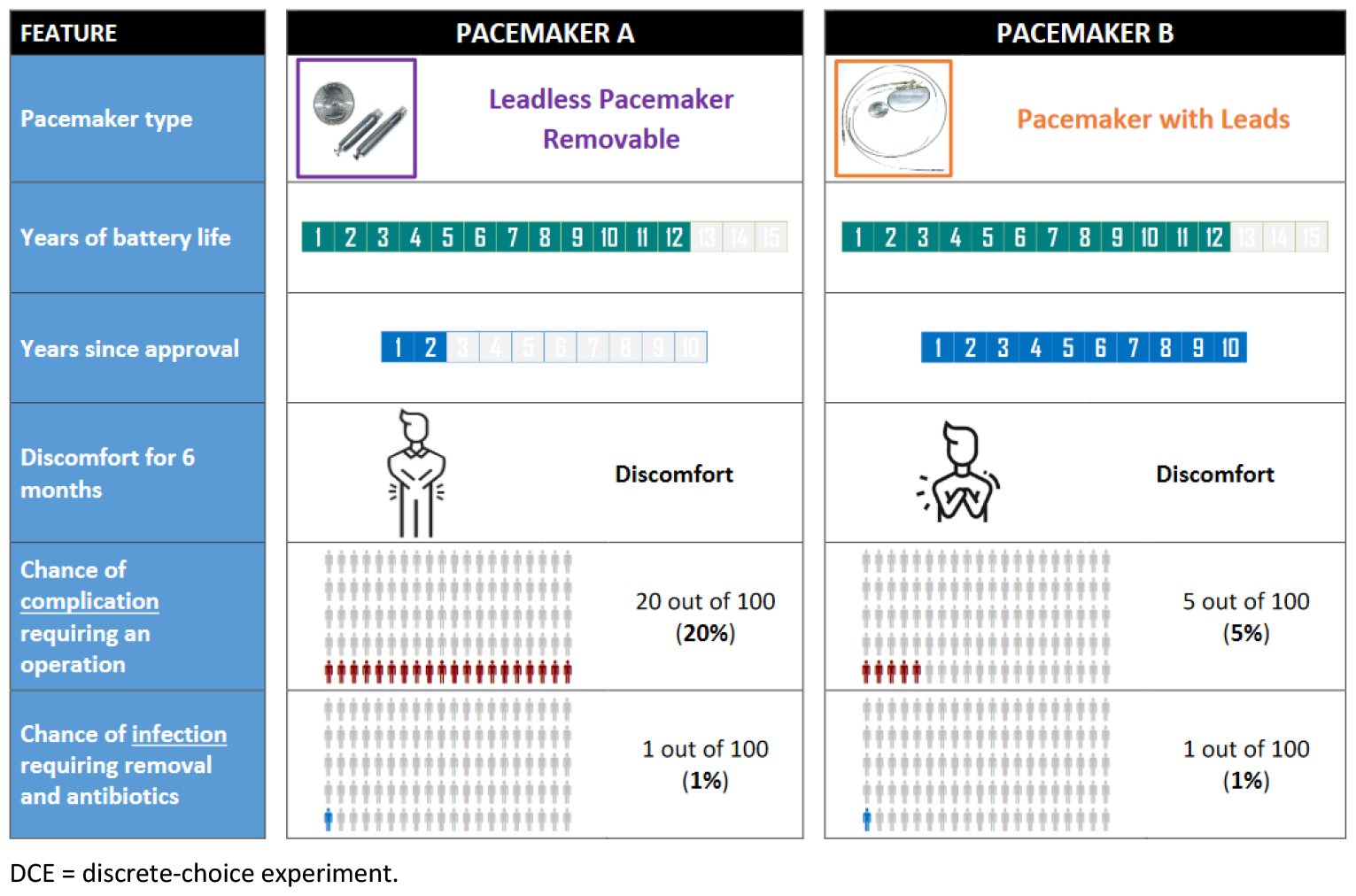
Example of DCE question. DCE = discrete-choice experiment.

**Figure 2.**
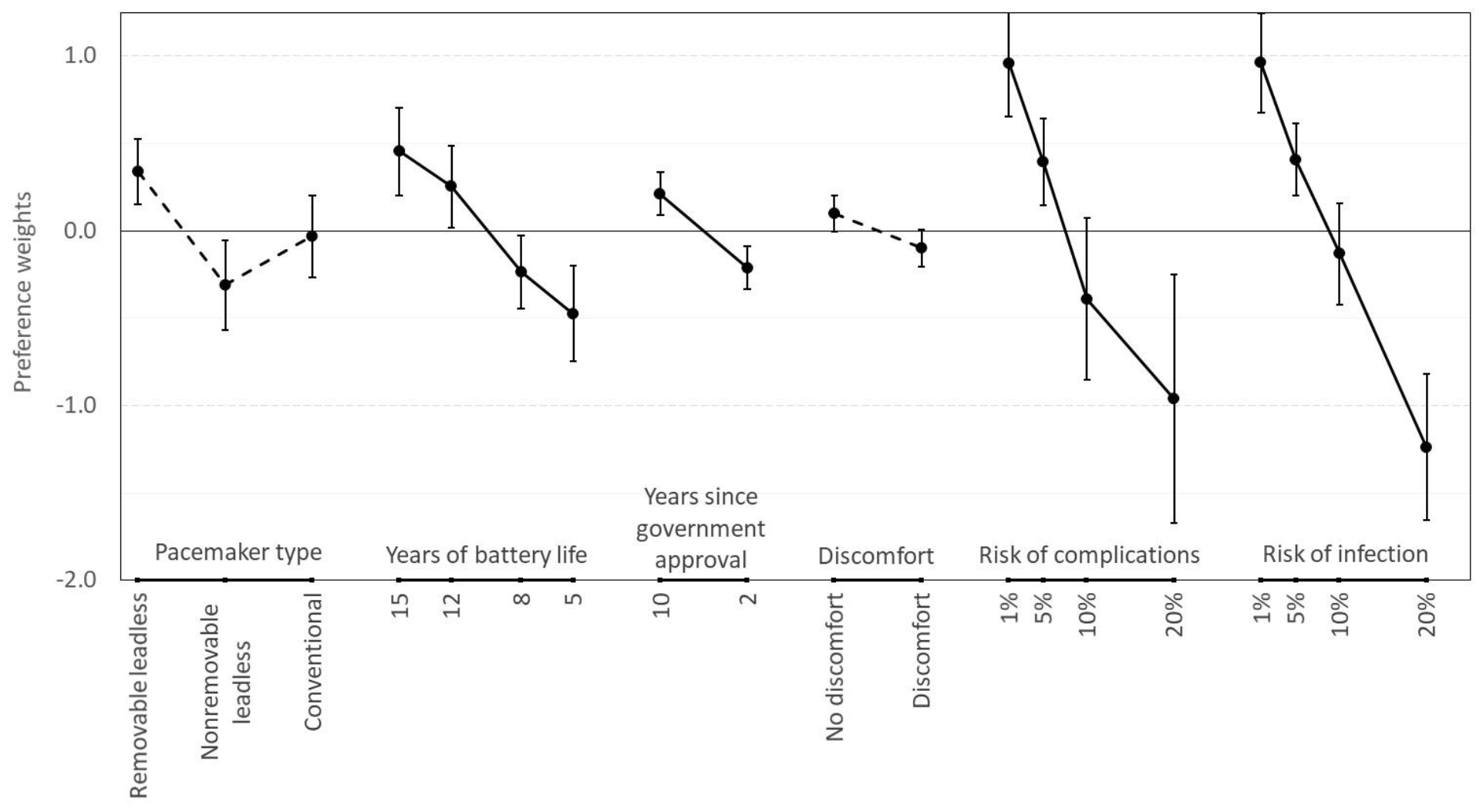
Error bars represents the 95% confidence intervals. Differences between categorical attribute levels are shown with dotted lines, and differences between continuous levels are shown with solid lines.

Preliminary LCA results indicated that a 2-class model provided the best fit for the data. Preference weights for the 2-class model for all attribute levels are shown in **Figure S1**. Preferences differed significantly between classes with regard to pacemaker type and years since regulatory approval (*p*< 0.001 for both), but not for the other attributes (*p*> 0.10 for all, **Table S2**). Therefore, the LCA model was re-specified to separately fit two classes of estimates for attributes representing pacemaker type and years since regulatory approval while constraining the coefficients for other attributes to be the same for both classes. As shown in **Figure 3 (Table S3)**, one latent class represented preferences favoring leadless pacemakers over conventional transvenous pacemakers (i.e. the leadless class), while the other latent class favored conventional transvenous pacemakers over leadless pacemakers (i.e. the conventional class). Participants with leadless-class preferences were insensitive to years since regulatory approval whereas participants with conventional-class preferences favored pacemaker alternatives with ten rather than two years since regulatory approval. Each latent class best represented preferences for about 50% of the sample. Participant-level class-membership probabilities for being in one or the other class were greater than 0.80 for 70% of participants, meaning that most individuals had a strong preference for one type of pacemaker.

**Figure 3.**
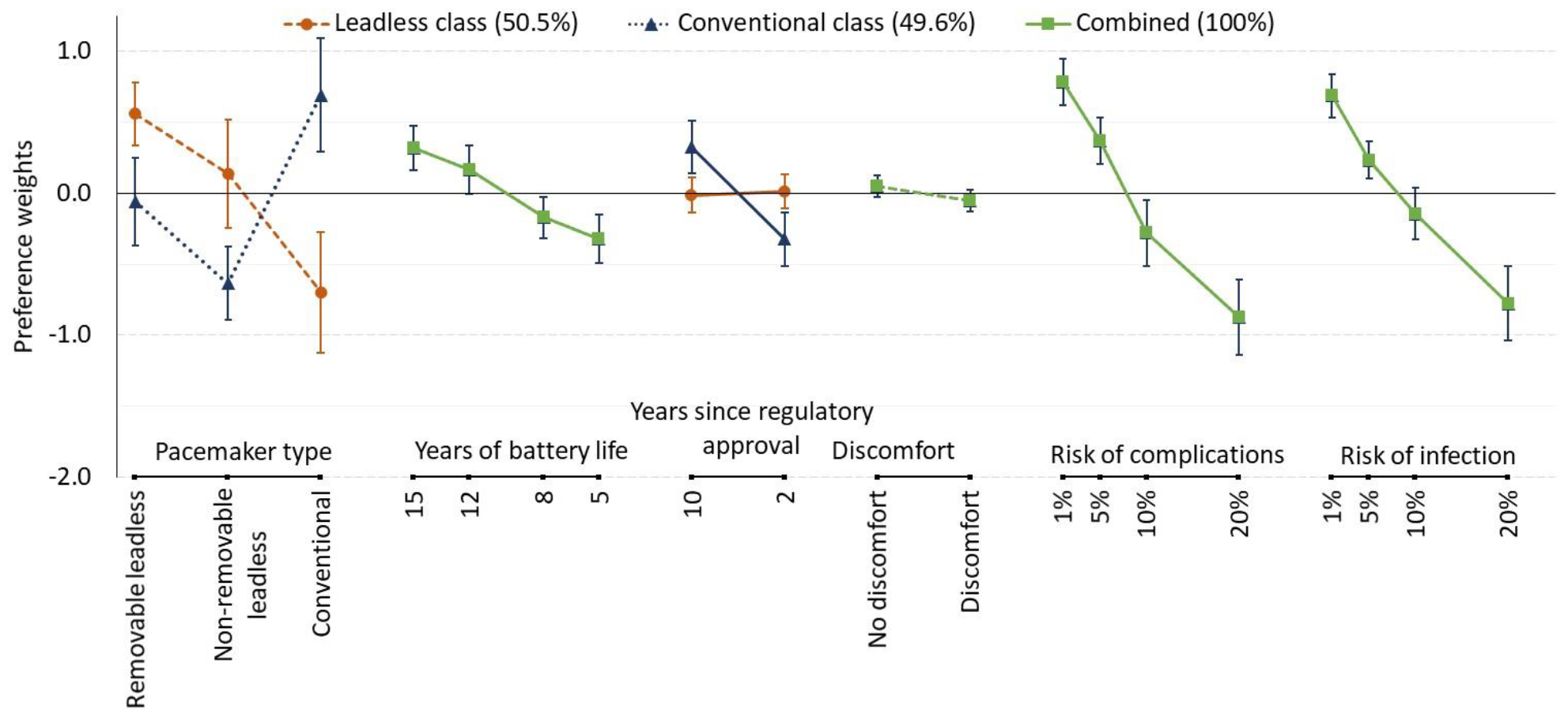
Error bars represents the 95% confidence intervals. Differences between categorical attribute levels are shown with dotted lines, and differences between continuous levels are shown with solid lines.

Participants with leadless-class preferences would accept up to a 13.4% risk of a complication or a 16.8% risk of infection to receive a removable leadless pacemaker instead of a conventional transvenous pacemaker, all else equal (**Table 2**). Conversely, participants with conventional class preferences would accept up to a 7.6% complication risk or an 8.9% infection risk to receive a conventional transvenous pacemaker instead of a removable leadless pacemaker. Both classes would accept about a 5% to 6% complication risk or a 5% to 7% infection risk, on average, to receive a removable rather than a non-removable leadless pacemaker.

**Table 2.**
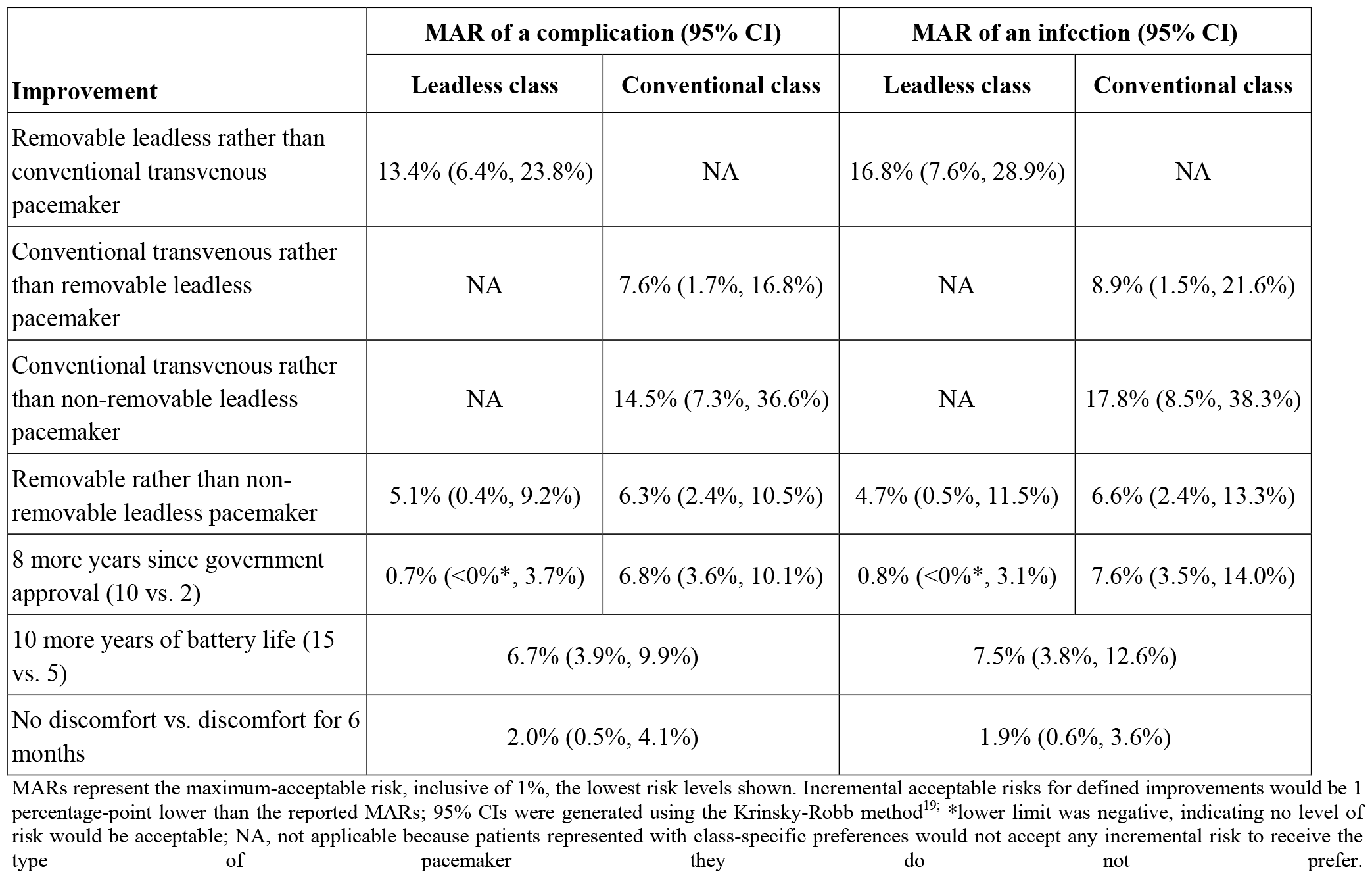
Maximum-acceptable risks (MAR) of complications or infections for specified improvements.

Participants with leadless-class preferences did not value the difference in time since regulatory approval; thus, they would accept no increased risk for pacemakers approved ten versus two years ago. Conversely, participants with conventional-class preferences would accept up to a 6.8% complication risk or a 7.6% infection risk to receive a pacemaker approved 10 years ago instead of 2 years ago. Both classes indicate acceptance of a 7% complication or infection risk to gain 10 years of battery life. On average, participants would only accept up to a 2% complication or infection risk to avoid discomfort associated with a pacemaker.

Three of the five participant characteristics included in the model were significantly associated with class membership. The following characteristics were independently associated with higher odds of membership in the leadless pacemaker class: lower BMI (OR: 1.03; 95% CI: 1.02-1.05), more active lifestyle (OR: 1.34; 95% CI: 1.08-1.64), and no history of major surgery within the past five years (OR: 1.52; 95% CI: 1.24-1.86). Female gender (OR: 1.13; 95% CI: 0.92-1.39) and age (per 10 years) (OR: 1.07; 95% CI: 1.15-0.99) were not significantly associated with class membership.

## Discussion

Accurately quantifying patient preferences for positive and negative aspects of medical devices can be valuable to various stakeholders. Medical device developers can use patient-preference information to aid product design.^20,21^ The United States Food and Drug Administration Center for Devices and Radiological Health recognizes the value of patient-preference data in regulatory decision making.^22^ In addition, such data could be of significant value to physicians and policy makers serving on patient safety advisory boards to guide external communications to providers and patients. ^23^ Patient-preference studies can also provide practicing clinicians a better understanding of acceptable benefit-risk tradeoffs from the patient perspective in the delivery of patient-centered care.

Our study is the first to quantify preferences for features differentiating leadless from conventional transvenous pacemakers. The most important features were the risk of a complication and the risk of an infection, both with risk levels ranging from 1% to 20%. On average, removable leadless pacemakers were the most preferred type of pacemaker over non-removable leadless pacemakers and conventional transvenous pacemakers. However, further analysis revealed that about half the participants preferred leadless pacemakers and half preferred conventional transvenous pacemakers. To obtain their preferred type of pacemaker, patients would accept increasing adverse-event risks from 1% to a maximum of 5% to 18%. These MAR estimates may seem surprisingly high. However, it is important to note that these estimates implicitly assume that the single adverse-event risk is the only downside associated with the pacemaker type, and a preference for a specific pacemaker type is the only advantage. If both risks are simultaneously relevant, estimated acceptance of each would be about half as large.^24^

There are few studies examining patient preferences pertaining to pacemakers. A study by Gulletta et al. reported that among patients who had received leadless pacemakers participating in a registry, patient preference was the factor driving the selection of a leadless pacemaker in 47% of patients 50 years or younger compared to only 6% of older patients.^25^ In our study, age was not significantly associated with leadless-class membership, but statistical power was limited with only 16 of survey participants reported being 50 years or younger. However, our study revealed that lower BMI, more active lifestyles and absence of major surgery in the past five years were independently associated with the leadless class. Individuals with lower BMI may prefer leadless pacemakers due to aesthetic concerns with pulse generators with conventional pacemakers, and individuals who exercise on a daily basis may prefer avoiding the longer period of physical activity restriction with conventional pacemakers. Regardless, these factors are insufficient to identify which types of patients prefer one pacemaker type over another. To ensure that patients receive a pacemaker that aligns with their preferences, they should be fully informed of the upsides and downsides of the various types of pacemakers available. Decision support tools that systematically provide this information in easy-to-understand language with useful graphics and elicit an individual patient’s preferences and values could help physicians to efficiently engage in shared decision making.

Although our study provides new insights about patients’ preferences relevant to choosing a pacemaker, a few limitations are relevant. First, our findings may not be generalizable to other samples. The age, gender and racial distributions were very similar in our study and the IDE study, but both samples under-represent minority populations, reflecting the common problem of minimal racial diversity in clinical research.^26^ Second, despite our concerted attempt to prepare participants to interpret pacemaker features as described, some may have assumed that risks are lower among pacemakers that have been on the market longer rather than considering the risk levels shown in DCE questions. Similarly, some may have assumed that longer time since regulatory approval was a positive indicator of greater dependability or provider experience. Others may have negatively viewed longer time since regulatory approval, perhaps representing a perception of outdated technology. Nevertheless, the data suggest that participants with preferences favoring leadless pacemakers may be representative of early technology adopters as they appeared to disregard time since regulatory approval. It is unknown whether participants had any knowledge about the IDE study or regulatory status for different types of pacemakers, but all were recruited before study results were published and before FDA approval of dual-chamber leadless pacemakers. Third, this study was designed and launched before the greater use of conduction system pacing. Therefore, the ability to achieve conduction system pacing with the conventional transvenous pacemaker and not with the leadless pacemaker, while currently clinically relevant, was not included as an attribute in the survey.

## Conclusion

Overall, our results indicate that patients have relatively strong preferences that align with removable leadless pacemakers or transvenous pacemakers, but there are other features that must jointly be considered after patients receive effective education about all pacemaker options and associated risks. Because patients place different levels of importance on pacemaker features, risks, and health outcomes, clinicians should take care to select devices in accordance with individual patients’ preferences.

## Data Availability

Individuals interested in accessing the study data are asked to contact the corresponding author with a written request, inclusive of a proposed study objective and analytic plan.

## Acknowledgments

We are grateful to the patient advisory group that provided valuable feedback throughout the study and the patients who devoted their time responding to the survey.

## Supplemental Material

Supplemental Methods

Figure S1

Tables S1-S3

Survey Instrument

